# Implementing Community-Based Blood Pressure Groups in Zimbabwe – findings from process evaluation of a pilot intervention

**DOI:** 10.64898/2026.03.05.26347087

**Authors:** Fadzaishe M. Mhino, Rudo M.S Chingono, Trevor Chivandire, Cuthbert Sekanevana, Chipo E. Mpandaguta, Takudzwa Mwanza, Alvern Mutengerere, Ardele Ndanga, Susana Scott, Pugie Chimberengwa, Justin Dixon, Chiratidzo Ndhlovu, Janet Seeley, Kalpana Sabapathy

## Abstract

**Background:** Hypertension is a significant health challenge in Zimbabwe and uncontrolled blood pressure (BP) highly prevalent. There are multiple barriers to optimal screening and management of BP and accessible and effective management strategies are urgently needed. We present findings from qualitative research on the potential for Community-BP (Com-BP) groups to address this gap.

**Methods:** The study was conducted in peri-urban (Chitungwiza) and rural (Chiweshe) areas in Zimbabwe. We aimed to assess, over three study phases, the feasibility and acceptability of Com-BP groups to facilitate self/peer measurement of BP, enable peer support and improve BP control.

We designed the Com-BP group model with key stakeholders before pilot implementation and evaluation. Focus group discussions, in-depth interviews and observations of group activities were conducted.

**Results:** Fourteen groups were established with a total of 151 community members (10-12 members per group). Digital BP machines were provided, a training manual for Facilitators and a Hypertension guide were produced to increase awareness about hypertension.

The intervention was found to be highly acceptable and feasible with the most important enablers identified as the provision of BP machines, joint facilitation by lay CHWs and peer facilitators (as community-champions) from within the community and peer support between members. Participants reported improved knowledge, medication adherence and sense of well-being.

**Conclusion:** The Com-BP group model enabled community members to actively take part in their own health-management and shows promise as an addition to facility-based care for BP control. Further research on sustainability, health system and health economic implications is needed.

## Introduction

### Community-based interventions to address hypertension

Hypertension is a leading risk factor for stroke, heart and kidney disease, sight-loss, and preventable reduction of disability-adjusted life years.(1,2) In Zimbabwe, most recent World Health Organisation (WHO) data indicate that 82% of adults aged 30-79 years with hypertension are inadequately controlled.(2) However, there are multiple health system and patient-related barriers including health-care worker attrition, resource shortages, and competing patient priorities (e.g. for work/income generation) so treatment remains sub-optimal, as elsewhere in much of Africa.(3,4) In public-health terms, cost-effective treatment options exist, but user fees and medicine costs borne by patients are prohibitive within the Zimbabwean context.(5) This is against the background of significant poverty with ∼50% of Zimbabweans living on <US$3/day, little access to health insurance, and inequality ranked among the highest in the world.(1,6) Lifestyle interventions to lower blood pressure (BP), early initiation onto treatment for those who need it, and greater efforts to improve treatment adherence are urgently needed in low income settings such as Zimbabwe to help lower the impact from the adverse cardiovascular outcomes caused by hypertension.

### Leveraging lessons from other conditions and geographical contexts

The WHO Global Report on Hypertension identifies community-based approaches to address hypertension as influential in the ability of some countries such as the Philippines and Bangladesh, to advance hypertension control.(2) Self-testing is widely used and recommended for many infectious conditions including HIV in SSA and more recently for COVID-19 testing worldwide.(7) Community Antiretroviral Therapy (ART) groups for people living with HIV, have become widespread in many southern African countries.(8,9) Evidence has grown on how groups help improve adherence and long-term treatment outcomes, reduce crowding within over-stretched health-facilities, and save costs for users and providers, particularly in rural settings where distance and access to healthcare facilities are disproportionately challenging.(10)

In the many settings, home BP self-monitoring is widely recommended to facilitate BP control.(11–13) Differentiated service delivery (DSD) has been embraced in South Africa, where integrated care for people living with chronic conditions including HIV and non-communicable diseases (NCDs) such as hypertension and diabetes aims to strengthen linkage, adherence and retention using a patient-centred approach.(14) These models enable individuals to participate in care planning, take greater ownership of their own health, and expand the successes of differentiated care models from HIV programming to other chronic conditions. They provide opportunities for translation of innovative approaches to develop interventions that address some of the unmet needs for BP control in Zimbabwe and similar settings.

The Community Blood Pressure (Com-BP) group study set out to examine whether community groups as an intervention could help address multiple steps in the cascade of care for hypertension – from screening and diagnosis to ongoing management, adherence, and retention in care – while empowering individuals to have greater control of their own health. The study aimed to pilot an intervention and generate evidence to inform a future study which would extend beyond hypertension to related co-morbidities, explore sustainability and examine integration into the local health system and scalability longer-term.

## Materials and methods

### Hypotheses

We hypothesised that Com-BP groups could contribute towards better hypertension care by: 1) increasing detection of hypertension by enabling community-based measurement at home or convenient community locations; 2) providing peer support for linking to care and improving adherence to clinical advice and treatment; 3) improving long-term optimal BP control by enabling ongoing regular self-monitoring and improved medication adherence.

### Objectives

The study was designed to address three key objectives: to produce, with input from key stakeholders (health service providers and users), a community group pilot intervention to improve BP control; to implement the pilot intervention and evaluate the appropriateness and feasibility of implementing groups, including identifying enabling factors, barriers, or unintended outcomes; and to interpret findings from the evaluation to define a future intervention that is scalable and sustainable for further research.

### Study design

Qualitative methods were used in the production and process-evaluation of the intervention, with additional quantitative surveys to describe the study population and evaluate key components, including changes in characteristics from baseline to endline.

### Study setting and population

The Com-BP study was conducted from October 2023 to April 2025, in peri-urban (Chitungwiza) and rural (Chiweshe) areas of Zimbabwe. Chitungwiza is a satellite town located approximately 25 kilometres from central Harare, the capital city of Zimbabwe. Chiweshe is a rural agricultural area in Mashonaland Central Province of Zimbabwe, North of Harare. At each site one health facility was purposively chosen to be the main point of referral for clinical care.

Nested within the Com-BP study was a project undertaken to explore acceptability and utility of information, education and communication (IEC) material in use by a local non-governmental organisation, SolidarMed, which is an implementing Partner of the Ministry of Health and Child Care (MoHCC) for improving access to NCDs. This formative work was based in rural primary health clinics supported by SolidarMed in three districts in Masvingo province.

The study population for intervention development in Phase 1 (Figure 1) comprised lay Community Health workers (CHW), healthcare stakeholders, study partners, and community members (health-facility attendees of varying age [adults ≥18y], purposively selected to balance gender and mix of health conditions, specifically to include some who were already living with hypertension). In Phase 2 the pilot intervention was conducted (May – October 2024) with community members who lived in the chosen sites and were expected to be available for the duration of the study, including CHWs who co-facilitated the groups. CHWs in Zimbabwe are lay members of the community, trained to facilitate community health programmes. Phase 3 involved key stakeholders (including health implementers, CHWs, nurses, and community members) from the same two communities and representatives from the MoHCC and local health leadership, to interpret study findings with the research team and make recommendations for future research and implementation.

**Figure 1:**
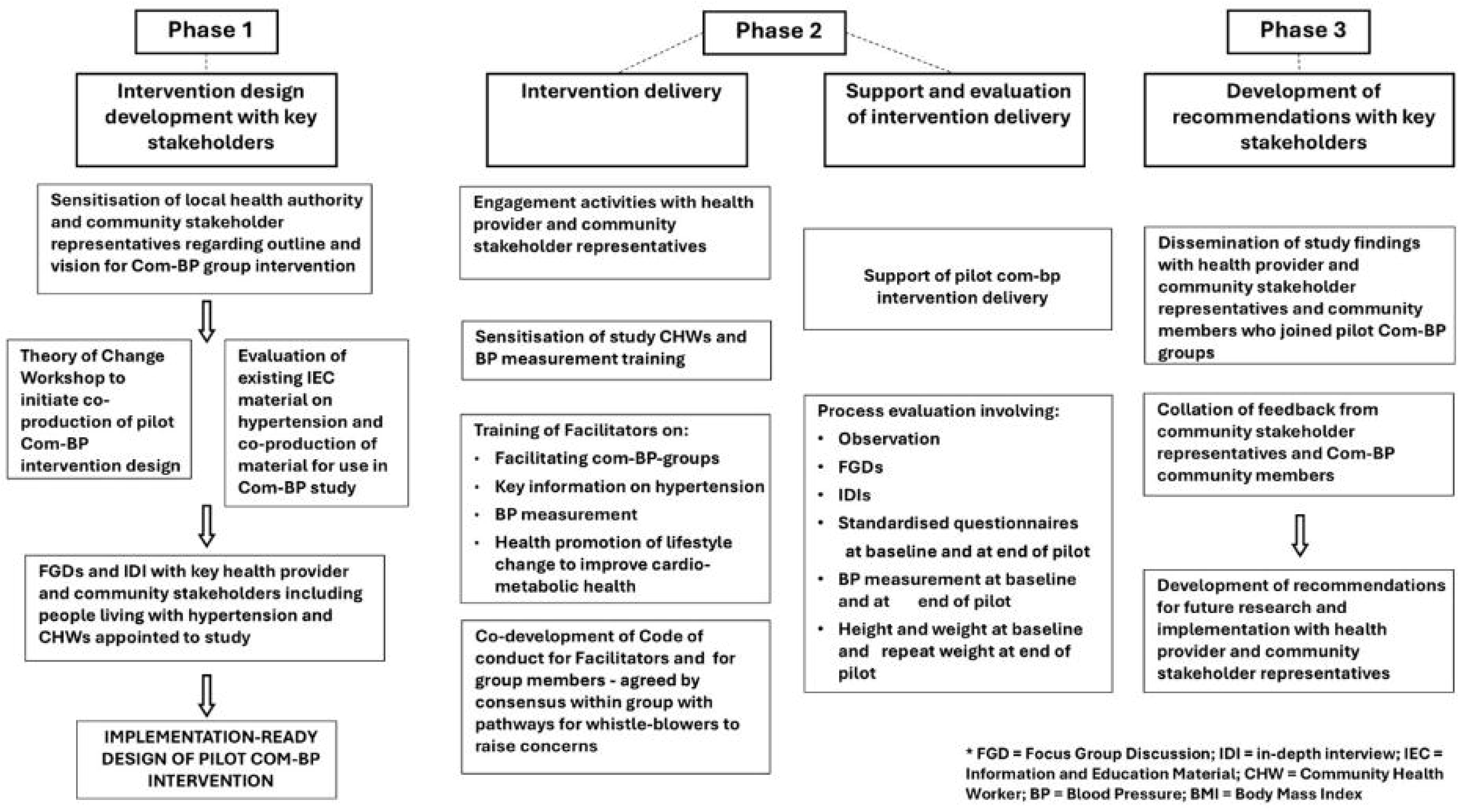
Phases of Research.

### Designing the Intervention

#### Phase 1

We began Phase 1 by exploring different perspectives on key themes related to hypertension. This involved focus group discussions (FGDs) with community members, stakeholders, and healthcare workers, and in-depth interviews (IDIs) with MoHCC representatives, local health leadership, clinic staff, and community members. A one-day Theory of Change Workshop was then conducted involving most of the same individuals for synthesis of ideas and collective discussion or “brainstorming”. Themes informed by published evidence on lessons learned from health-related community groups in Africa, were developed by the study team to guide for further exploration with stakeholders. The themes included pre-existing hypertension service provision and uptake; priorities and strategies for improving hypertension care; the perceived appropriateness and feasibility of Com-BP groups; the structure, delivery options, and requirements for Com-BP group models; potential synergies of Com-BP groups with other existing initiatives for hypertension; and potential barriers and enablers for optimal implementation (Table 1). By the end of the Workshop consensus was achieved among stakeholders present, on the necessary components of pilot Com-BP groups and an implementation strategy was developed for piloting and evaluation in Phase 2.

**Table 1:**
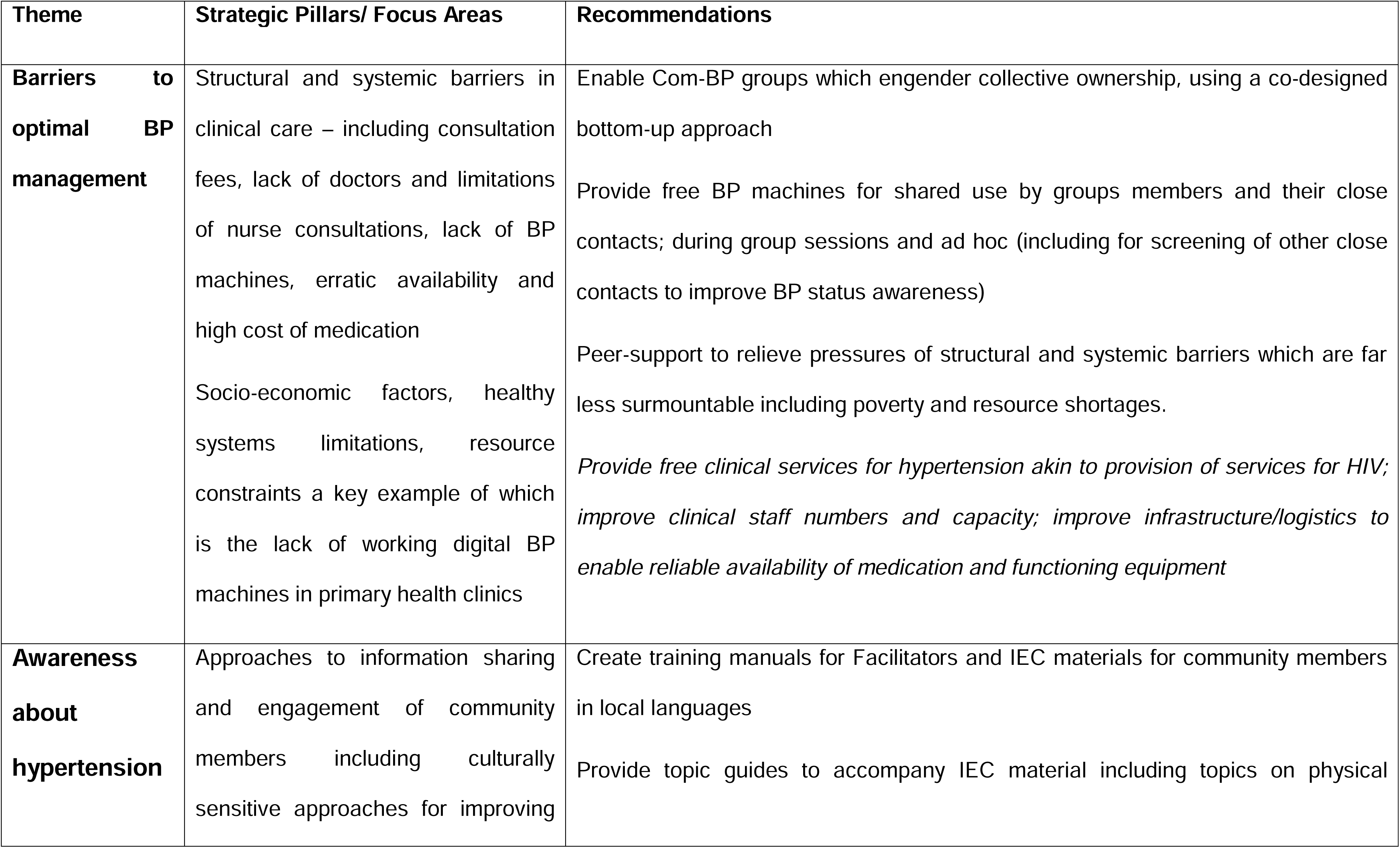
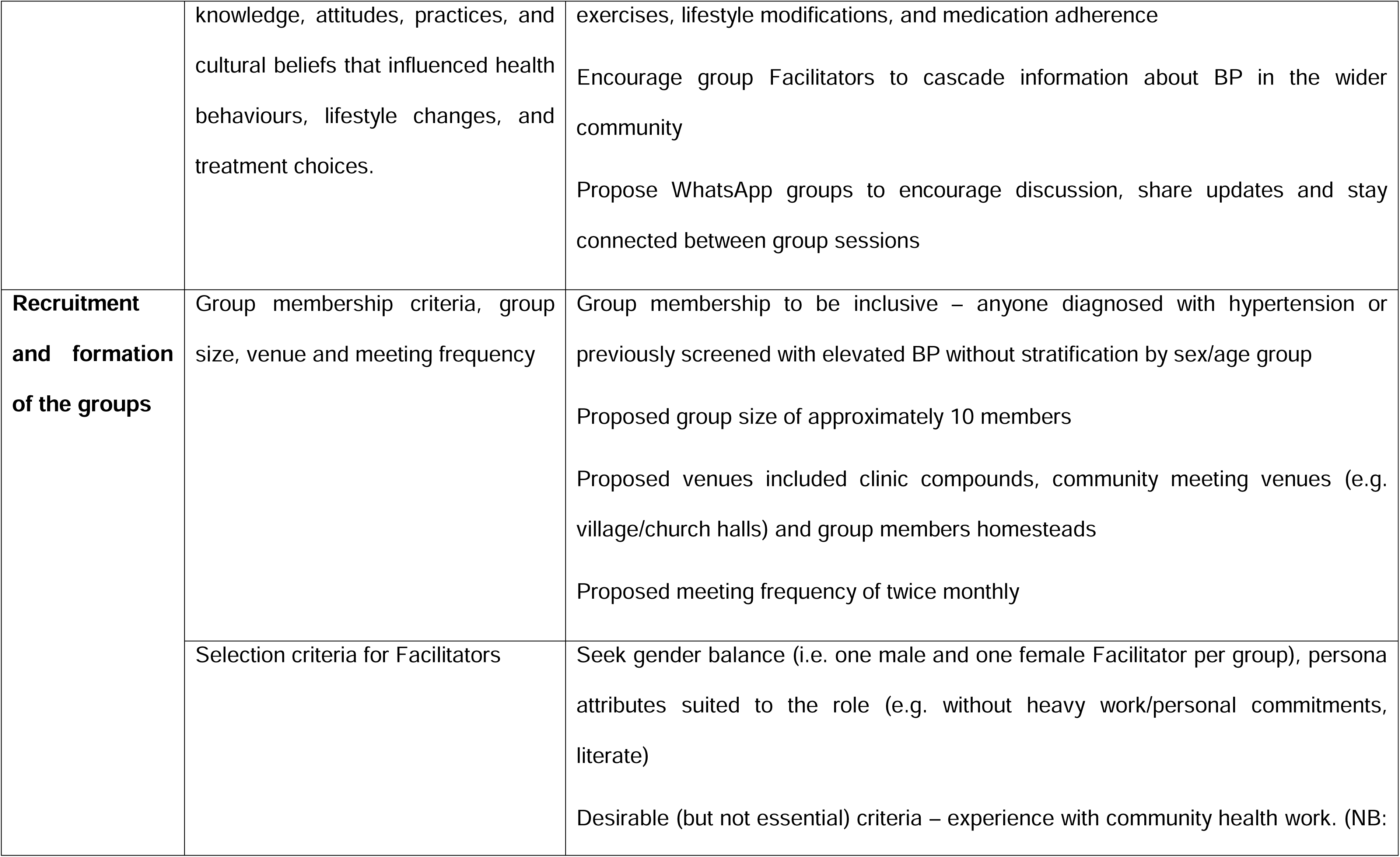
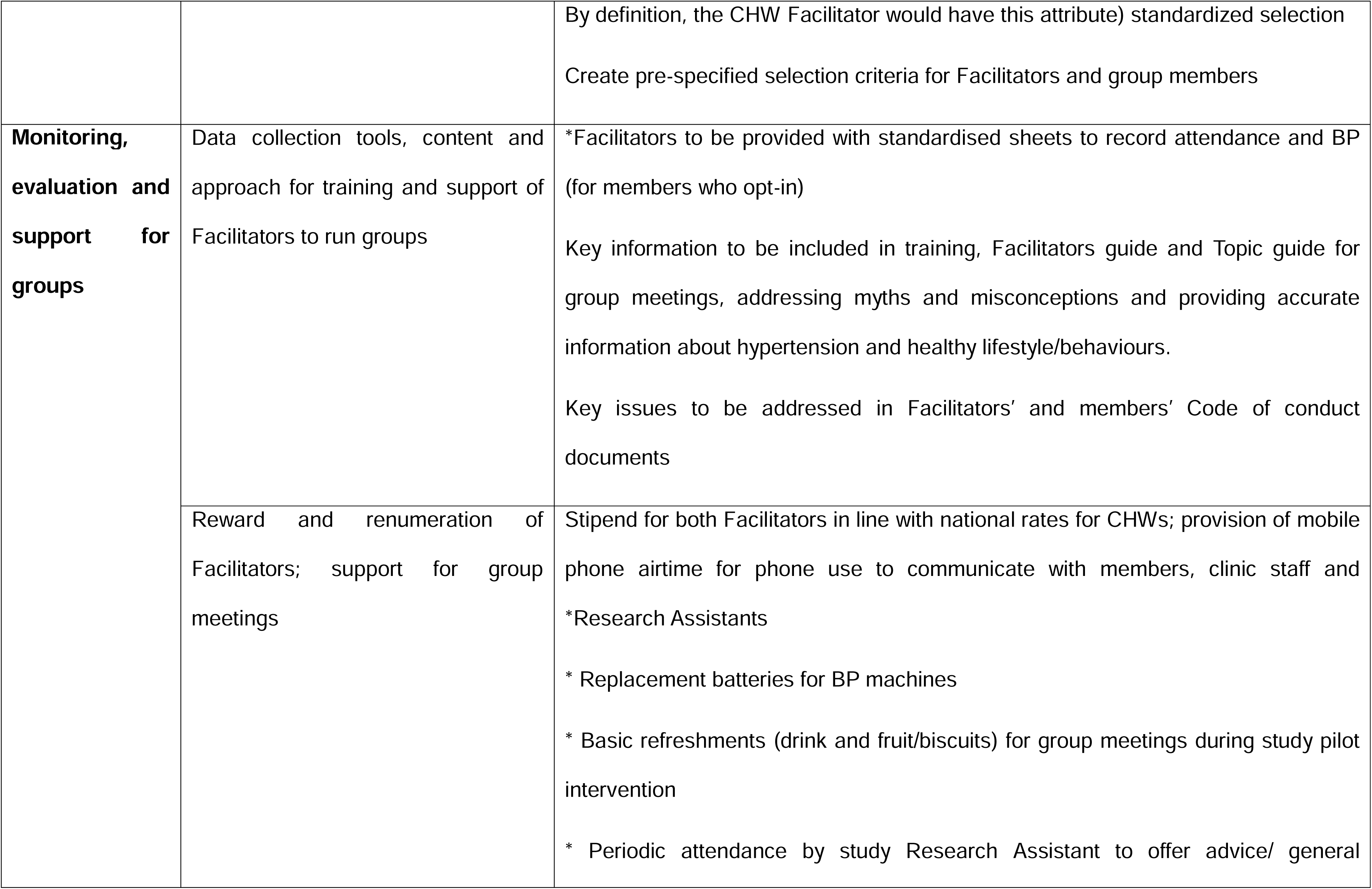
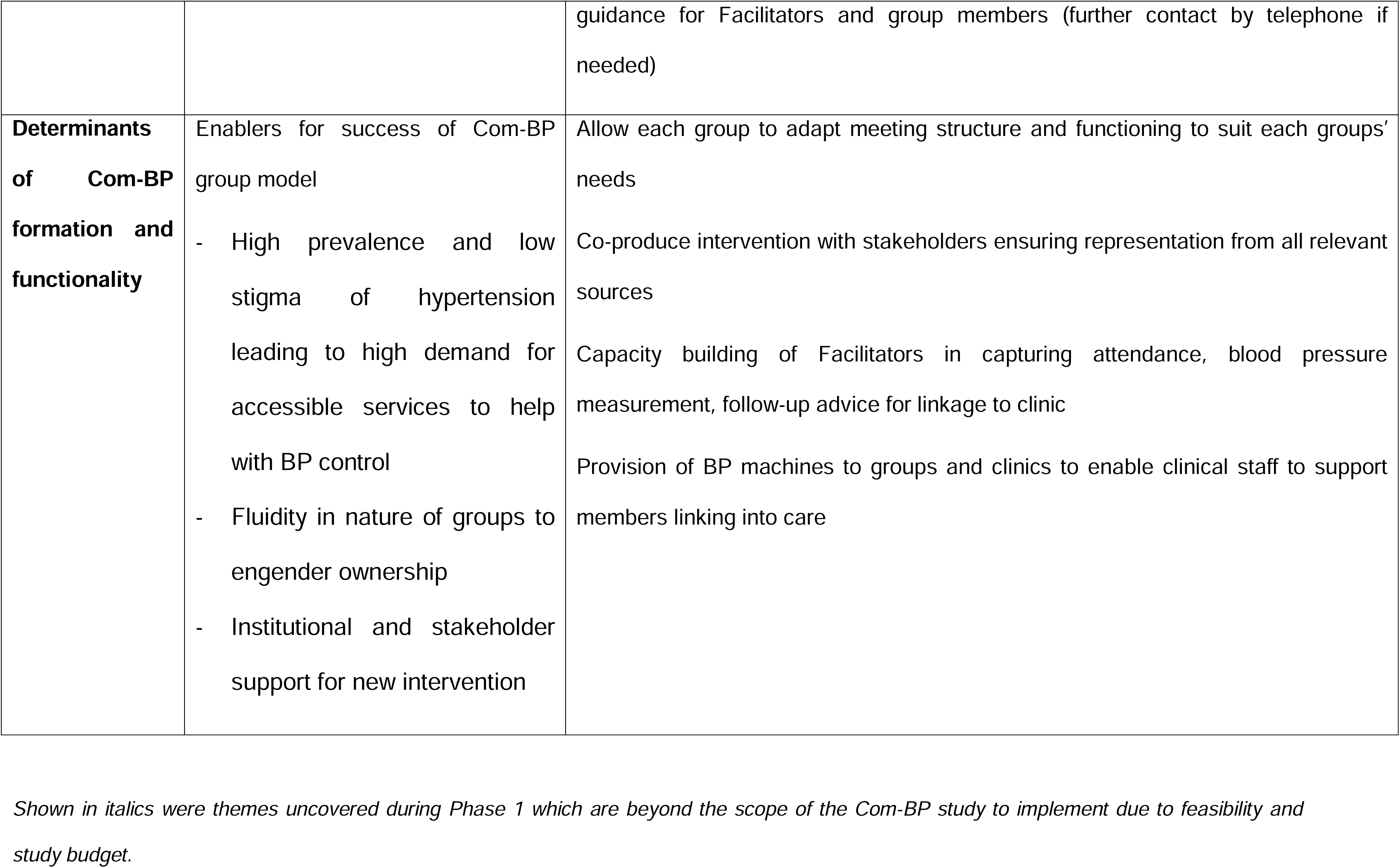
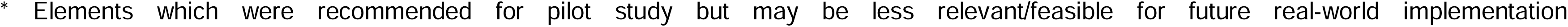
Summary of Phase 1 findings informing the COM-BP intervention design.

The study team developed a context specific hypertension information booklet based on pre-existing content on hypertension for lay persons from the WHO (<u>Hypertension</u>), British Heart Foundation (<u>High blood pressure (Hypertension) - causes and symptoms - BHF</u>) and content used in Zimbabwe by SolidarMed (S1_File). In addition, a group facilitators manual and a guide to Understanding Hypertension film was developed to address knowledge gaps and needs shared by community members and stakeholders during Phase 1. A code of conduct for Facilitators and group members was also produced.

#### Phase 2

Pilot Com-BP groups were implemented, guided by formative research in Phase 1 as listed in Recommendations for pilot intervention of Table 1.

A process evaluation with the pilot groups was conducted using qualitative research methods guided by the MRC Process Evaluation Framework and the Consolidated Framework for Implementation Research (CFIR).(15–17) IDIs, participatory workshops, and observations were conducted guided by a standardised tool of key themes. For both the IDIs and workshops, purposive sampling of group members was done to ensure broad representation of group members (for example, age and gender balanced). Key themes, guided by the MRC framework and explored in the research topic guides, included fidelity to the draft intervention design, feasibility of delivery, acceptability and perceived quality of services, and social harms/unintended consequences. Observation of selected Com-BP groups meetings were conducted by research assistants, using a structured observation guide (S2_File) that captured intervention fidelity, acceptability, and determinants to the successful implementation of Com-BP groups. Each group was observed at least once a month, and detailed observation notes were taken by a trained Research Assistant. Attendance and retention were monitored using attendance records kept by Facilitators, who also documented BP measures done during group meetings.

A quantitative survey, using a standardized questionnaire, was administered to all Com-BP group members at the first meeting and last study-supported meeting approximately 5 months later. Socio-demographic characteristics and knowledge, attitudes, and practices (KAP) about hypertension were examined. Three BP measures were also taken at both time points by Research Assistants trained to comply with WHO guidance for BP measurement.(18) Acceptability of the Com-BP groups was also examined in the follow-up survey. Findings from the quantitative survey and BP measurement are fully described in another manuscript by Mhino/Ndanga *et al*.(19)

#### Phase 3

Findings and lessons learned from Phase 2 were examined in Phase 3 to evaluate the scope for Com-BP groups as a potentially viable intervention to be implemented at scale. MoHCC representatives and other key stakeholders involved in Phase 1 research were presented with evidence from process evaluation of the pilot, during a one-day Study Findings Dissemination Workshop. A summary of findings was presented and material produced during Phase 1 and 2 of the study (see Results) were shared, before being submitted to MoHCC for review ahead of widescale dissemination.

### Data management and analysis

All interviews and FGDs were transcribed and translated from Shona to English. Observations were written up into detailed field notes. All the qualitative data were manually coded on the electronic version of each document in Microsoft Word. Thematic analysis, following Braun and Clarke’s six step process(20), was used to analyse data collected as part of the evaluation. We used both a deductive and an inductive approach to coding.(20,21) Initially a coding framework was deductively established based on existing literature on community peer to peer groups supporting chronic conditions (such as community groups for people living with HIV), the CFIR, and MRC Process Evaluation Framework. Other emerging themes were then coded inductively. To ensure rigour and shared understanding of the coding framework, the research team applied the initial codebook to a subset of the data (three IDIs, two FGDs, and five observation notes). Once consensus on the codebook was reached within research team members, the remaining dataset was analysed. Emerging patterns and differences were identified, and interpretations were based on triangulation across data sources and team discussions.

Quantitative descriptive analysis of participant characteristics, KAP and medication adherence scores, BP and BMI is described in the manuscript by Mhino/Ndanga *et al*.(19)

### Ethics

Permission to conduct the study was obtained from the Biomedical Research and Training Institute ethics board, London School of Hygiene and Tropical Medicine (Ref: 29785), Medical Research Council of Zimbabwe (MRCZ/A/3069), and Research Council of Zimbabwe. Approval was granted by relevant authorities in Zimbabwe, including the Ministry of Health and Child Care (MoHCC), the Chitungwiza City Health Department, the Chitungwiza District Administrator, and Howard Mission Hospital in Chiweshe. All participants provided informed consent and all those who could not write were assisted by a nominated witness.

## Results

During Phase 1, a total of 24 IDIs were conducted; with healthcare stakeholders (n=10), and community members (n=14). Four FGDs (two in Chitungwiza and two in Masvingo) were conducted with 12-14 community members. In Phase 2, four FGDs (n12-14) and 25 IDIs were conducted with Com-BP group members (n=15) and key informants (n=10). A total of 71 group observations were conducted across the 14 groups (an average of 5 observations per group). IDIs were for approximately one hour, FGDs for approximately three hours and observations for the duration of the relevant groups’ sessions, typically 1-2 hours.

### Design of pilot Com-BP group intervention

The COM-BP intervention was designed to be aligned with the Recommendations in Table 1 arrived at after research conducted in Phase 1 with heath provider and community representative stakeholders. Key components of implementation (Figure 2) included provision of content on health education, use of existing community structures (CHWs and systems to enable linkage into care), and tailoring of group meetings as fitting for the respective groups, determined collectively by members themselves. While most groups were not stratified by age/sex, one dedicated group for older community members (≥60 years) was formed, as an extension of the existing structure facilitated by the Zimbabwe Older People’s Association for community group meetings. Access to BP monitors was managed by the CHW, and the nurse in charge at the local primary health clinic (PHC) was the primary custodian of the BP machines. Residents were able to measure their BP for free, facilitated by the Com-BP group facilitators, even if they were not part of the group. Referral for further care was to be made by the CHWs. Facilitators were paid a small stipend aligned with the MoHCC rates for renumerating CHWs in Zimbabwe.

**Figure 2:**
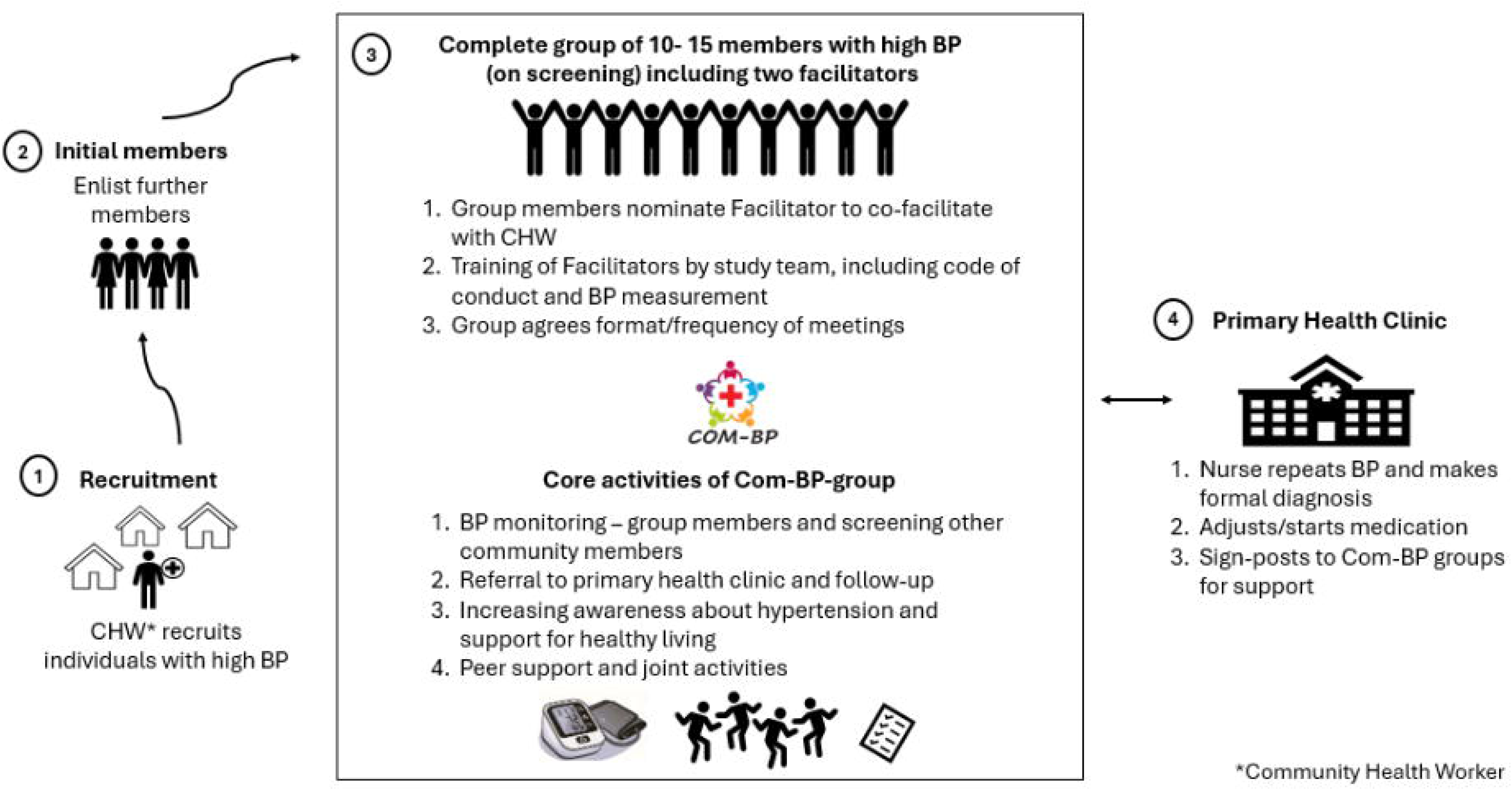
Design of the Com-BP pilot intervention.

Fourteen CHWs (one per community) were first selected from those who served the health facilities within those areas. To set up the groups, each CHW recruited a handful of already known hypertensive adults or those recently screened with high BP (Figure 2). Further members were drawn in by word of mouth. CHWs screened interested community members with the BP machines provided and referred those with high BP to the clinic. Individuals who were confirmed as hypertensive at the clinic were eligible to join. Once 10 members were identified a group was considered complete, however, up to five more members were allowed to join during the course of the pilot.

CHWs screened interested community members with the BP machines provided, to identify those with high BP, who were not previously diagnosed. Individuals were recruited to the group once diagnosis of hypertension (systolic and/or diastolic BP ≥140/90 mmHg) was confirmed at the clinic. Once 10 members were identified, a group was considered complete, allowing for up to 5 more members to join.

Fourteen groups with 10 individuals per group (including two Facilitators) were formed, seven in Chitungwiza and seven in Chiweshe, respectively. While the target was for 140 Com-BP group members, 150 were recruited in total with one member being lost to follow-up by the end of the pilot period (approximately 5 months). Members were predominantly female (N=137, 79%) and ages ranged from 30-92 (average 54y). BP monitors were donated to the groups and batteries replaced by the study team during the pilot intervention period.

Two Facilitators (CHW and group member) were assigned to each group to distribute responsibility and facilitate peer support between them. The CHW was the primary Facilitator and a second Facilitator to co-facilitate the group was chosen by members. The second Facilitator had to meet the inclusion criteria (Table 1), namely, be literate, have the time to volunteer, and be willing to adhere to the Code of Conduct for Facilitators which detailed requirements for confidentiality, respect and other relevant principles of Good Clinical Practice (S3_File). Facilitators received a monthly volunteer stipend and training on basic information regarding hypertension, including proper use of BP monitors, interpretation of readings, and protocols for repeat measurements. They were also trained, to promote healthy lifestyles, using materials from the World Health Organization (WHO), such as videos and visual aids.

The research team trained the Facilitators and supported them in forming the groups. Group members then determined the frequency and location of their meetings by consensus within the group. Most groups held meetings weekly for the first month, settling to fortnightly thereafter. Meeting places ranged from group members home compounds, town halls or informal locations (e.g. under a specific large shady tree in the rural areas). Study research assistants also attended the first two meetings to provide support and supervision on BP measurements, use of digital BP machines, and how to use content from the Com-BP Facilitators guide and Guide to Understanding Hypertension documents (S4_File and S5_File respectively), which were developed in Phase 1 with input from key stakeholders, to stimulate group discussions. Subsequent telephone contact was maintained when Facilitators needed any advice.

Sessions were dynamic, incorporating innovative health education methods based on the content provided by the study team in the Facilitator’s guide and guide to Understanding Hypertension documents. For example, if healthy diet was the chosen topic, Facilitators would relate information on that topic from the Facilitator’s guide and Guide to Understanding Hypertension and members would discuss previous misconceptions and reflect on opportunities and barriers to making improvements in the future.

### Unpacking the Com-BP group intervention

Six key themes emerged from Phase 1 research for intervention development and Phase 2 process evaluation of pilot Com-BP groups (Table 2). These themes include community understanding of hypertension; health-seeking behaviour and treatment experience; acceptability and feasibility; impact; sustainability of the Com-BP groups; and recommendations for the future from community members’ perspectives. These themes are discussed in detail below.

**Table 2:** Thematic areas identified during observations, FGDs and IDIs during Phase 1 and 2 and illustrative quotes and observations.

| Theme |  |
| --- | --- |
| **1. Community understanding of hypertension** |  |
| a. Evil spirits cause hypertension and its adverse effects | “So when you then try to tell someone how BP works [contributes to strokes] as you speak with them they will argue with you that it is not BP but some evil spiritual attack.”<br>(Female, 50-54 years) |
| b. Hypertension only affects elderly people<br>c. Lack of awareness that treatment interruptions are harmful | “In 2013, I was diagnosed with BP. Then I was on medication for about two and a half years. Then I stopped the medication when I realized (*sic*) I was better. I stopped on my own because I looked at my age and I didn’t think it was good that I would be taking BP medication.” (Male, 35-39y). |
| d. Low treatment literacy regarding different medications and doses which need tailoring for individual patients | A woman (80-84y) reported that when she ran out of her own medication, which was Nifedipine, she started taking her brother’s pills also for hypertension, not realising that there may be differences. The CHW who tested her blood pressure during a group session when she was unwell noticed that it was not Nifedipine and this may have been the reason she was experiencing side effects. (Observation 03, Site 2, Group 9) |
| **2. Health-seeking behaviours and treatment experiences** |  |
| a. Financial barriers to adherence | “...The other reason for me not to visit the health facility is that sometimes I do not have the money for medical fees” (Male, 45-49y).<br>‘I defaulted for three months due to lack of money to purchase the drugs...’ (Female, 70-74y) |
| b. Use of traditional remedies | Group members confessed to using both traditional herbs and prescribed medication concurrently, at times or substituting the latter for the former, when they did not have money to purchase their BP meds. (Observation 03, Site 2, Group 8) |
| c. Use of medication bought ad hoc in pharmacies without prescription | “Most of the time I go to the pharmacy and get tested. But I don’t go often, I only go when I feel that today my body is a bit weak.” (Male, 50-54y). |
| **3. Acceptability and feasibility of the Com-BP intervention** |  |
| a. Need for accessible interventions to help address hypertension | “From my perspective, people accepted and loved the program because most times people would have to go to the hospital to get their BP measured but recently it has been easy for them as they can have their BP checked at any time within the community...<br>People do not have the money to go to the hospital to get their BPs checked so they accepted this initiative and everyone within the community including those who are not in |
|  | the groups are getting their BPs checked.” (Female, 50-54y) |
| b. Easy access to BP machine | “The groups had an impact, especially because of the BP machines. If one has repeated high BP readings after regular checks, they are given a referral letter to go to the hospital for medical assistance” (Female, 50-54y)<br>“Others from the community are excited about the intervention. They are coming for BP checks...The visitors who come are measured BP as well (sic). They know the days when we will be having BP checks and the days for the sessions as well.” (Male, 70-74y) |
| c. Peer support | “I have seen that the groups are helping because we are socialising and interacting well. We are now like one family. We listen to each other’s problems, and we comfort each other, and we can help each other when we see that one of us has high BP... So those are some of the things that are helping as we meet. It is moulding us, helping us to socialize better with others than before” (Male, 50-54y)<br>A notable observation was that when a group member experienced a problem, peers within the group were supportive and sought to offer emotional comfort and practical advice, even when the problem was not directly related to BP and involved challenges of |
|  | daily life (Observation 04, Site 1, Group 7). |
| d. Shared ownership and commitment to group's collective success | In one instance, despite a facilitator's late arrival, participants remained engaged and it was observed that members were not restless, but maintaining high spirits and curiosity about how the session would benefit them. (Observation 01, Site 1, Group 6). |
| **4. Impact of Com-BP intervention** |  |
| a. Management of BP | "...I am seeing a change because my BP no longer gets higher like what it used to be like 140/100 and above. It's now better and it now ranges from 128/80." (Female, 30-34y).<br>Similar reports were noted during group observations, for example when one participant explained that her BP has improved, from measures as high as 168/101mmHg to 141/74mmHg on the day of the observation (Observation 05, Site 2, Group 14). |
| b. Improved linkage with clinical care | "I was given a referral slip which I took to the Central Hospital. I went and saw the doctor and he added more pills. Since taking the pills they added, my BP now is good it is now low [within normal range]. I am taking the pills two in the morning and one at night. So, it [the referral] really helped me." (Female, 50-54y). |
| c. Improved adherence to treatment | Participants reported adherence to medication and commitment not to "default". |
| and medical advice | (Observation 03, Site 1, Group 5). |
| d. Improved lifestyle and behaviours | The group influenced me to change my lifestyle as a hypertensive patient, I stopped many things because I used to eat fried foods and do just anything, I did not exercise but learnt that I should exercise, eating boiled food, food with little oil instead of fried food my lifestyle and social life changed because I got the knowledge from the sessions so it helped me. My lifestyle changed because before joining the Com-BP group I used to eat too much salt and fatty foods, did not exercise, and would be lazy telling myself that it's for youngsters I realised that exercising is also beneficial to older people like me who have BP."( Female, 55-59y) |
| e. Scope for reduction of adverse outcomes caused by hypertension | "Some had strokes and did not know it was BP. Others in the community are now in wheelchairs because of BP. So, it's helping, strokes will be reduced as people gain access to regular BP checks."(Female, 50-54y) |
| f. Scope to improve other medical conditions and multimorbidity | "They noticed that my diabetes and BP are no longer high....(*helped by*) what I learned from the group." (Group member, Site 1, Female, 35-39 years).<br>"...because sometimes I think that in life I am useless and there is nothing I can do but if you get words of encouragement it will help in knowing that you can make it" (Female, 30- |
|  | 34y). |
| **5. Sustainability of Com-BP groups** |  |
| a. On-going group meetings | *Reported during interactive session in Phase 3 Study Findings Dissemination Workshop* |
| b. Formation of new groups | *Reported during interactive session in Phase 3 Study Findings Dissemination Workshop* |
| **6. Community perspectives on future implementation** |  |
| a. Call for increase in number and scope of Com-BP-groups | "I think more sessions should be added... adding other topics that we have not learnt from before. Adding other topics that you think can be useful to our health like sessions on HIV, Diabetes and heart problems" (Female, 65-69y) |
| b. Routine monitoring of BP by CHWs | *Reported during interactive session in Phase 3 Study Findings Dissemination Workshop* |
| c. Free consultation and medication for hypertension | *Reported during interactive session in Phase 3 Study Findings Dissemination Workshop* |

#### 1. Community understanding of hypertension

Although many individuals were aware of the existence of hypertension, there were significant knowledge gaps among community members in both the rural and urban sites. Most participants demonstrated limited knowledge on what causes hypertension, leading to varied awareness regarding what factors contribute to hypertension. While some participants were aware it is caused by excessive salt intake or ageing, others attributed hypertension to being caused by stress alone. Belief in misinformation was common, and many individuals believed that strokes (which are an important consequence of uncontrolled hypertension) are caused by evil spirits (Table 2, 1a).

The belief that hypertension is a condition which only affects older adults, and that younger people could not be affected was also uncovered. It was one identified cause for failure to adhere to treatment advice and treatment interruption (Table 2, 1b/c). The commonest medicines reportedly prescribed were Nifedipine, Hydrochlorothiazide (HCT) and Atenolol but participants were often unaware that there are different drugs to treat hypertension. There was poor understanding that drugs are not interchangeable and that doses are tailored to individual patient needs, resulting in medication sharing, under- or over-dosing, and other unsafe practices (Table 2, 1d). This is compounded by the fact that patients do not have any patient held medical records to refer to or show to other health providers such as pharmacists or clinicians in a different clinic.

#### 2. Health-seeking behaviours and treatment experiences

Most group members, regardless of whether they lived in rural or urban areas, expressed that they occasionally miss medication doses due to the associated costs, including consultation fees and the cost of medications (Table 2, 2a). Participants reported that on average the cost of the medication was less than US$5 which is a significant proportion of average household income in these communities and could even be as high as US$15. A significant challenge in hypertension management is the impact of financial constraints on medication adherence and appropriate treatment, often compounded by the poor treatment literacy described earlier.

Community members use a variety of approaches to manage their hypertension. While many participants reported using prescribed medications, some opted for alternative remedies, such as various herbal treatments, and others combined both methods. Economic hardships often led some individuals to discontinue their prescribed medications and rely solely on alternative options due to cost considerations. Many group members were convinced that traditional herbal remedies are effective at treating hypertension and a key benefit was that it was a cheaper option than prescribed medication (Table 2, 2b).

IDIs conducted at both sites revealed that most individuals visited health facilities infrequently for routine check-ups. Many individuals reported that they find it more convenient to visit pharmacies for BP checks, primarily due to the lower consultation fees. Even when feeling unwell, individuals often described their symptoms to the pharmacist and obtained medications over the counter, bypassing formal medical consultations (Table 2, 2c).

#### 3. Acceptability and feasibility of the Com-BP Intervention

The community group model was perceived to be highly feasible and community members strongly welcomed the idea of the Com-BP group intervention considering gaps in existing services (Table 2, 3a). The sessions were well attended and individuals invited others to join, leading to the formation of groups that included couples and family members. As interest grew, some groups expanded to more than 10 members. The additional members joined during the course of the study, having heard positive remarks from founding group members.

The easy access to BP machines was an important reason why individuals welcomed the intervention both for group members, as well as other community members who were also able to access the machines (Table 2, 3b). A culture of peer support was clearly evident, with much mutual encouragement to stay committed to the groups and to their health. As members learned about hypertension during group sessions, they shared personal testimonies to demystify myths and misconceptions. Culturally relevant messages to enhance engagement and achieve understanding was used. In one instance, participants planned and performed a show for other members illustrating the consequences of untreated hypertension with a drama about a family who believed BP was caused by evil spirits.

The strong peer support was also identified as a helpful means of dealing with stresses of daily life commonly reported by members (Table 2, 3c). In addition, participants exhibited strong commitment, camaraderie, and a shared sense of purpose and ownership, with regular attendance, punctuality and social support between members (Table 2, 3d). The final study supported group session was observed to be a very emotional experience for group members.

#### 4. Impact of the Com-BP Intervention

BP lowering was observed in the quantitative analysis comparing BP measures done at baseline and in follow-up surveys (detailed in study by Mhino/Ndanga *et al*).(19) Community members also perceived this to be a key impact of the intervention as they frequently observed their BP measures lowering over time (Table 2, 4a). BP monitoring within the groups, coupled with the follow-up and advice from CHW/facilitators, appeared to reinforce adherence to medication and lifestyle changes. Individuals who had previously defaulted on treatment re-engaged with care. As the primary facilitators were trained CHWs, they were familiar with navigating health system protocols and able to provide referral slips to the local primary health clinic when needed (Table 2, 4b).

Group members benefitted from improved knowledge about both the causes and consequences of hypertension, and the importance of adherence to medical advice and prescribed treatment (Table 2, 4c) Additionally, behaviour changes and lifestyle modifications including healthier eating, exercise, and increased social interaction were adopted (Table 2, 4d) and groups engaged in dance and exercise during meetings. There was optimism that support for lowering BP could lead to reduced adverse outcomes, such as strokes (Table 2, 4e).

Participants also reported improvements in their other medical conditions such as diabetes and mental health (Table 2, 4f).

#### 5. Potential sustainability of Com-BP groups

Feedback from dissemination meetings held after the program’s conclusion during the final phase of the study suggested there was some indication of potential long-term sustainability of the intervention due to the enduring nature of the groups, albeit for the relatively short duration following the pilot study. Of the 14 groups that were created during the study, members present from 10 of the groups reported that they were still meeting at least once a month (at the time of the meeting nine months after the end of the study pilot). This was enabled in part by the BP machines donated to the groups by the study, with group members making contributions for purchase of batteries to power the machines. The group of older adults not only continued their own meetings but had also successfully supported the development of a second older adults’ group and was preparing to establish a third group. Of the four groups that were no longer meeting, one shared a plan to begin monthly contributions, with the goal of enabling each member to procure their own BP machine which could support BP measurement and monitoring among their peer networks, beyond the Com-BP group. Another group reported that their machine was not working and this was a key reason for discontinuation of the group.

Across both sites, clinic nurses continued to provide support when needed. In turn, they referred newly diagnosed patients seen in the facilities to groups, for support and monitoring of BP. Further, CHWs who were the primary facilitators of the Com-BP groups chose to initiate BP checks when they were conducting community maternal health checks. Others took screening to community gatherings, referring those with elevated BP measures to clinic.

#### 6. Community member and health stakeholder perspectives on future implementation

There was a call for increasing the number of groups or having larger numbers within each group to widen the reach of the intervention. Requests were also expressed for increasing the number of BP monitoring machines available for use by the groups and increasing the topics for discussion during sessions, including on other non-communicable diseases (NCDs) such as diabetes and mental health (Table 2, 6a). The latter included a request to integrate a counselling component for greater mental health support within groups.

Community members advocated for increased access to BP monitoring by MoHCC, asking that CHWs be routinely provided with BP machines to use in the community. In addition, they asked for referral pathways to PHCs which included free consultation and medication for hypertension. Analogies were drawn with initiatives for addressing HIV including a call for assistance for people living with hypertension in the same way as for people living with HIV, namely with consultations and drugs.

Stakeholders involved in developing the pilot intervention were presented with a briefing document (S6_File) and a short film about the study (S7_File). Feedback was collated as recommendations for future research and implementation.

## Discussion

The Com-BP model was found to be an acceptable and feasible intervention, with a significant sense of ownership and positive impact perceived by community members involved in the pilot. The intervention’s design and implementation were effective, with three defining contributors to success: strategic provision of BP machines to groups for use in the community; the peer led facilitation by nominated group members and trusted local CHWs; and grass-roots involvement of members in deciding the nature of conducting groups.

Community health programmes which focus on NCDs are not widely delivered despite significant need and potential scope for them to have meaningful contribution to patient well-being.(22) Consistent with our findings, existing evidence suggests that community-based interventions can improve knowledge on risk factors and influence physical activity levels and dietary practices which improve NCDs.(23) A recent systematic review examined community-based models of care for hypertension in the African region and found that there was insufficient evidence to refute or recommend individual models.(24) The review identified two studies (in Kenya and Nigeria) which examined community club models to help hypertensive patients with adherence. However, these did not include screening of new patients to improve detection of hypertension, or had used health-care workers to perform BP measurement without the same level of “peer-hood” as was the case in our model. (25,26) In our Com-BP group model there was co-facilitation by a peer CHW and group member nominated by the group members themselves. A study from Matabeleland South Province in Zimbabwe explored approaches to improve hypertension management at community and primary care levels, but BP machines were not provided for community members to freely access when requested.(27)

Donation of BP machines with replacement batteries when needed, was an invaluable factor in facilitating regular BP monitoring in the current study. This component directly addressed a major barrier in the Zimbabwean context and health system - that of logistical and financial costs associated with accessing routine monitoring at public health facilities.(28) Members of Com-BP groups and additional community members were able to get their BP checked for free, a service they would normally have to pay for at a PHC or pharmacy. A further financial disincentive is that health facility attendance often involves loss of income from time spent waiting in over-crowded clinics. Ready access to BP monitoring allowed the early detection of high BP, for individuals who would otherwise most likely have only sought health services after developing sequalae of chronic uncontrolled hypertension or worse with acute presentations such as strokes.

The involvement of both CHWs and community members as facilitators was instrumental, given the shared background and experience of peers from the same community, with the benefit of not being received as top-down intervention. With some basic training, group facilitators were able to play a crucial role in participant recruitment, facilitation of group meetings, linkage to care and adherence support. Their deep-rooted connections within the community fostered trust and open engagement. This approach was highly effective, particularly in engaging men who are typically harder to reach in health programs.(29) In addition, the intervention’s reach beyond group members created a peer-driven ripple effect highlighting the intervention’s ability to address a health need which was strongly felt as a gap in the community. Task sharing and task shifting, through training CHWs or other allied health professionals to deliver basic health care, have been previously documented as effective strategies.(3,30,31) The facilitators’ role in offering health advice and referral to clinic when needed bridged a key gap in the existing health system, where follow-up care is often inconsistent.(28) This holistic approach is crucial for managing a chronic condition like hypertension.(3)

Having peers with lived experience was an important driver for others’ motivation to improve their health outcomes. Many individuals had first-hand exposure of the adverse effects of uncontrolled hypertension, reinforcing their commitment to BP monitoring. Further the motivation and commitment exhibited by group facilitators acting as “community-champions” towards raising awareness, promoting lifestyle changes and providing adherence support to group members also motivated individual members to control their BP. Group cohesion and morale also played a pivotal role in the seeming sustainability of the intervention.

Underpinning these successes was the design of the intervention with detailed experienced and invested stakeholders, and a “bottom-up” approach to grassroots shaping of how each group tailored the intervention to their needs. Members within each group actively contributed to shaping their sessions, fostering a sense of collective ownership. By alleviating some of the psychosocial burdens that can impede treatment adherence, the intervention created a more resilient and supportive environment to maintain well-being. The provision of IEC material with approaches for lifestyle modification tailored for the setting, enabled demystification of cultural and religious myths and correction of misconceptions about hypertension which are highly prevalent,(32,33) replacing them with accurate health information instead.

Sustainability of peer-based models is an important consideration for long-term relevance.(34) There is some indication that the Com-BP group intervention could be sustainable based on the feedback received during the Phase 3 Study Findings Dissemination Workshop. The ability to continue meeting and/or peer support to establish new groups with collective funding and procurement of further BP machines, demonstrates a shift from participants as beneficiaries and passive recipients, to that of active agents with responsibility for their own health. This demonstration of collective responsibility or “ubuntu” and self-sufficiency showcases the ability to deliver a member-driven model. This is compelling evidence for the potential success of the Com-BP group intervention in helping to create sustainable health practices that are deeply embedded within a community’s social capital.

### Strengths and limitations

There was overwhelming support and endorsement of the Com-BP intervention as described earlier. A key strength of the design is the hybrid approach which combines elements of both implementation science and clinical evaluation. This allowed the researchers to assess the realities of living with hypertension, the intervention’s acceptability and feasibility, and understanding of the factors which enabled success. In addition, we also gained insights into the potential impact on health outcomes described in another publication by Mhino/Ndanga *et al*.(19)

However, there were also some limitations to consider, the first of which is because of the nature of recruitment into the groups. By definition, only those who were interested in being involved in community groups chose to join. It follows that the Com-BP group model and its positive effects should be viewed as an additional option to facilitate improved health outcomes for those who are amenable, rather than a substitute for universal individualised care, as community groups may not be an attractive option to certain sections of the community. Further, the observed improvements of health outcomes and behaviours are indicative but not conclusive, and benefits were self-reported and liable to desirability bias. Also of note is that the study provided support for group meetings, albeit with modest provision of soft drinks and biscuits.

While the Com-BP group intervention did include active male participation, there were notably fewer men and those involved were generally older men. This is consistent with other evidence that men’s participation in community programs is challenging.(29) The pilot nature of the study also meant that there were only a small number of groups, supported for a limited duration. Despite the program’s effectiveness, some participants faced financial barriers that hindered their access to healthcare and limited the full potential of the benefits from early detection and referral achieved by Com-BP groups. The relatively modest budget of the study constrained the degree of support which could be provided, although this was also a strength of the real-world possibilities and what is achievable with limited resources.

Other concerns which have been highlighted by studies involving community groups include the possibility of inadequate moderation and risk of misinformation or inaccurate content being shared or reinforced without adequate qualified professional support or oversight.(35,36) This highlights the importance of training and supervision, clear codes of conduct for facilitators and group members, and pathways for involvement by qualified professionals, as was implemented in the Com-BP study. Inequitable access is another concern, especially for minoritised groups who may be marginalised due to disabilities or language barriers, for example.(22)

### Impact and recommendations for the future

The briefing document we have produced summarises key elements for others seeking to deliver Com-BP groups in similar settings and other study resources have been made available to MoHCC and local and regional stakeholders.

Due to the high prevalence of hypertension in southern Africa, hypertension was the priority area and starting point for exploring proof-of-concept of using community groups for NCDs. In future, there is scope to extend the model to address other chronic conditions including diabetes. In South Africa, the Central Chronic Medicines Dispensing and Distribution programme has been implemented to improve access to medication collection by providing community-based drug collection points. This could be another component for further exploration in Zimbabwe and similar settings, as part of differentiated service delivery, alongside single-pill combination treatment to improve BP control, reduce pill-burden, and enhance efficiency.(14,37)

The feasibility of scaling up the intervention, sustainability over extended periods and extension to multiple related long-term conditions including diabetes with detailed evaluation of clinical outcomes, were called for by health stakeholders involved with the study. Cost-effectiveness analyses and health systems research are also key to fully understand how the intervention could be incorporated into existing health services, not least to identify efficiency gains from being able to transfer some health services away from heavily burdened health facilities.

Our study presents a strong case for the feasibility and acceptability of a community-based BP intervention in Zimbabwe and similar low-resource settings, but also any health system seeking to incorporate strong community involvement and ownership of health and well-being.

## Acknowledgements

We acknowledge the contribution of all key stakeholders involved in design of the intervention and especially thank community members for their participation in the research.

## Data Availability

The datasets generated and analysed during the Com-BP study are available from the corresponding author upon reasonable request. Supplementary materials S1_File, S4_File, S5_File, S6_File, and S7_File contain identifiable images of participants and study personnel (with informed consent) are available to be shared by the corresponding author upon reasonable request.

## Supplementary Files

1. S1_File: SolidarMed Hypertension Information Leaflet - *Available from collaborating author upon request*
2. S2_File: Com-BP Session Observation Guide
3. S3_File: Code of Conduct for Com-BP Facilitators
4. S4_File: A Facilitator’s Guide - *Available from collaborating author upon request*
5. S5_File: Understanding Hypertension - *Available from collaborating author upon request*
6. S6_File: Developing an intervention to improve blood pressure control in Zimbabwe (The Com-BP study) – Executive Summary - *Available from collaborating author upon request*
7. S7_File: Com-BP Study Film - *Available from collaborating author upon request*

