## Supporting File 2 for "Implementing Community-Based Blood Pressure Groups in Zimbabwe – findings from process evaluation of a pilot intervention"

**SUPPORTING INFORMATION 2: Com-BP Session Observation Guide**

| |
|---|
| **Location of observation** |
| **Date of observation** |
| **Start and end time of observation** |
| **Name of group** |
| **Session number** |
| **Name of facilitator** |
| **Name of researcher observing** |
| **Date of writing up notes** |
| **Materials used to deliver the session** |

**1. General impressions** *(What is your general impression of the observation? Include the degree to which the intervention was implemented as intended)*

______________________________________________________________________

______________________________________________________________________

______________________________________________________________________

______________________________________________________________________

**2. Setting**

*- Describe the community, including buildings, space*

*- How does the space feel?*

*- What is the set-up of the group session, tables, where people are sitting, standing or moving?*

*- How comfortable is the setting?*

*- How does the weather impact the setting?*

*- Did the setting change during the period of observation? If so, how?*

*- Any other details of the setting*

______________________________________________________________________

______________________________________________________________________

______________________________________________________________________

______________________________________________________________________

**3. People and interactions**

*- Who is present during the observation (group attendance= n/N)?*

*- Any visitors*

*- Describe the group dynamics (What were the interactions between different people? Did this change over time? Were there any differences in interaction between males/ females, different ages?*

______________________________________________________________________

______________________________________________________________________

______________________________________________________________________

______________________________________________________________________

**4. Processes**

*- Describe the BP measuring process*

*- Describe the capturing of registers process*

*- Describe the exercising process*

*- Include in each description details such as what happened, length of time, how providers and other group members engaged*

**5. Group Discussion**

*- Topic of the day*

*- Were there any questions raised?*

*- Was the group participative?*

______________________________________________________________________

______________________________________________________________________

______________________________________________________________________

______________________________________________________________________

**6. Delivery and receipt of services**

*- Were there any adaptations to the planned delivery of services?*

*- How did the facilitators appear to feel about facilitation?*

*- How did the group members appear to feel about engaging in the different activities?*

*- Were there any aspects of the delivery that could have been improved?*

______________________________________________________________________

______________________________________________________________________

______________________________________________________________________

______________________________________________________________________

**7. Informal conversations**

*- Did you have any informal conversations with anyone present? If so, with whom? Describe each interaction separately, including where and how you spoke, and what was discussed (include direct quotes).*

______________________________________________________________________

______________________________________________________________________

**8. Reflection**

*- How do you think your presence influenced what happened during the observation?*

*- Are there any particular things that you would like to explore in more detail in further data collection?*

*- Is there any part of the observation that made you feel uncomfortable or uneasy?*

*- Are there any ethical challenges that were encountered in this observation?*

*- Are there any changes to the tool that you would suggest based on this observation?*
