## Supporting File 3 for "Implementing Community-Based Blood Pressure Groups in Zimbabwe – findings from process evaluation of a pilot intervention"

**SUPPORTING INFORMATION 3: Code of Conduct for Com-BP Facilitators**

**Background**

The document details the terms of reference of a Facilitator for the Com-BP Study conducted on behalf of the Biomedical Research and Training Institute (BRTI).

**Position Summary**

- To report to the BRTI staff assigned to the group. It will be the responsibility of the group members to select their Facilitator. This decision will not be influenced by the study team or local leadership, except within the predetermined selection criteria and terms of reference.
- The Facilitator is responsible for:
    - building relationships with community members and organizations
    - mobilizing group members in the Com-BP research activities.
    - facilitating group meetings,
    - safe keeping and sharing of BP machines,
    - follow up of group members (CHW),
    - filling in of registers and debriefing of group proceedings to the study team as per training guidelines.
    - The Focal Person also works to ensure that the Com-BP research project's activities are conducted in a way that is respectful and of good clinical practices and community values, and priorities.

**Facilitator Criteria:**

1. A resident of the particular community.
2. 18 years and above
3. Responsible and respected member of the community
4. Someone who can read and write
5. Known hypertensive/recently diagnosed/Community Health Worker (CHW)

**Responsibilities:**

1. Booking and setting up of the meeting place
2. Notification of local leaders/gatekeepers of meeting dates
3. Follow up on members with reminders for the meeting
4. Ensure safe keeping of all BP machines, keeping a register to track loans and returns of machines
5. Ensure that all necessary registers are completed at the expected timelines
6. Ensure safekeeping and accountability of refreshments for the group

7. Give feedback (via regular phone calls within 2 days after a meeting) to the study team on what transpired during the group meetings.

**Requirements:**

1. Excellent verbal and written communication skills
2. Ability to work with limited supervision and take their own initiative
3. Willingness to walk around the community to locate potential and follow-up participants.
4. Ensure that you maintain and respect group confidentiality

**Stipend:**

The Com-BP Group Study team will provide a volunteer stipend of US$25/month as a way to acknowledge and appreciate the work that has been done by the Facilitator. This stipend will be provided subject to the conduct laid out in this document.

**Signing of the Code of Conduct**

- I have understood and agree to the above. These have been clearly explained to me
- I agree to be a Facilitator for the duration of the Com-BP Group Study, subject to consensus within the group and study team
- I agree to maintain and respect the confidentiality of group members and the group code of conduct and values
- I agree my position is not that of authority, but primarily to facilitate and support the group
- I agree that the study team and/or Biomedical Research and Training Institute is not responsible for any misconduct on my part within and outside the groups

Name of Facilitator (please print): Signature: Date:

Name of Witness (please print): Signature: Date:

Name of study team representative: Signature: Date:
